# Multiplexed, Microscale, Microarray-based Serological Assay for Antibodies Against All Human-Relevant Coronaviruses

**DOI:** 10.1101/2020.09.03.20179598

**Authors:** Erica D. Dawson, Laura R. Kuck, Rebecca H. Blair, Amber W. Taylor, Evan Toth, Vijaya Knight, Kathy L. Rowlen

**Affiliations:** InDevR Inc. 2100 Central Ave., Suite 106, Boulder, CO 80301, USA; Children’s Hospital of Colorado, 13123 E 16th Ave, Aurora, CO 80045, USA

**Keywords:** COVID-19, SARS-CoV-2, human coronavirus, serology, immunoassay, VaxArray

## Abstract

Rapid, sensitive, and precise multiplexed assays for serological analysis during candidate COVID-19 vaccine development would streamline clinical trials. The VaxArray Coronavirus (CoV) SeroAssay quantifies IgG antibody binding to 9 pandemic, potentially pandemic, and endemic human CoV spike antigens in 2 hours with automated results analysis. IgG antibodies in serum bind to the CoV spike protein capture antigens printed in a microarray format and are labeled with a fluorescent anti-species IgG secondary label. The assay demonstrated excellent lower limits of quantification ranging from 0.3 – 2.0 ng/mL and linear dynamic ranges of 76 to 911-fold. Average precision of 11% CV and accuracy (% recovery) of 92.5% over all capture antigens were achieved over 216 replicates representing 3 days and 3 microarray lots. Clinical performance on 263 human serum samples (132 SARS-CoV-2 negatives and 131 positives based on donor-matched RT-PCR and/or date of collection) produced 98.5% PPA (sensitivity) and 100% NPA (specificity).

## 1. INTRODUCTION

A novel coronavirus (nCoV) was first identified in December 2019 in Wuhan, China, and was declared a worldwide pandemic by the World Health Organization on March 11, 2020. Coronaviruses infecting humans belong to either the alpha or beta coronaviruses. 229E and NL63 are alphacoronaviruses, while coronaviruses HKU1, OC43, SARS-CoV-1 (the virus causing Severe Acute Respiratory Syndrome that first circulated in 2003), MERS-CoV (the virus causing Middle East Respiratory Syndrome), and the current SARS-CoV-2 virus causing COVID-19, formerly referred to as nCoV, belong to the betacoronaviruses.^1,2^ While 229E, HKU1, OC43, and NL63, known to continually circulate in humans, generally produce mild symptoms, MERS-CoV, SARS-CoV-1, and SARS-CoV-2 more recently have crossed over from animal reservoirs into humans^1,2^ and can produce symptoms that are quite severe. Moreover, SARS-CoV-2 has been shown to effectively transmit person-to-person and cause significant morbidity and mortality, as evidenced by the over 5.4 million US and 21.9 million worldwide confirmed cases of the virus as of August 18, 2020 and the associated 3.5% global mortality rate.^3^

The role of serological testing to measure antibodies to SARS-CoV-2 has been the subject of recent debate,^4,5^ given that we do not yet fully understand the antibody levels required for seroprotection or how long protective antibodies may persist. In addition, the Food and Drug Administration’s waiver of the requirement for Emergency Use Authorization (EUA) for SARS-CoV-2 serological assays as long as internal validation has been conducted and appropriate limitations language are included in product labeling^6^ resulted in a large number of serological tests introduced into the market for diagnostic use. As of August 18, 2020, 397 commercialized antibody detection immunoassays are listed on FIND’s website.^7^ and numerous reports of serological assays with variable performance have been recently highlighted.^8-11^

Regardless of the utility and role of SARS-CoV-2 serology assays for diagnostic and seroprevalence applications, a critical application of serological testing for SARS-CoV-2 is monitoring antibody response to COVID-19 vaccine candidates during pre-clinical and clinical trials to enable a full understanding of the immune response post-vaccination.^4,12^ There are currently 202 SARS-CoV-2 candidate vaccines in pre-clinical or clinical development.^13,14^ Binding assays including ELISAs are an alternative to the gold standard virus neutralization assays, as they do not require cell culture or significant biosafety containment measures, are straightforward to conduct, and have been shown to correlate with virus neutralization assays for other coronaviruses.^15^ In addition, antigen binding assays currently commercially available or in clinical use typically establish specificity using pre-pandemic serum samples which cannot conclusively show that pre-existing antibodies to the endemic coronaviruses do not cross react with SARS-CoV-2.

Serological testing that assesses binding to a variety of coronaviruses is important for: (1) screening enrollees before trial admission to establish baseline antibody titers, (2) monitoring longitudinal profiles of antibody response post-vaccination, and (3) comparing pre- and post-vaccination immune responses in pediatric vs. adult cohorts. Recent reports and workshops have highlighted the unknowns about pre-existing immunity to SARS-CoV-2.^16,17^ In light of these unknowns and recent discussion around the potential for antibody dependent enhancement,^18-20^ monitoring the serological responses before and after immunization to other human coronaviruses, such as SARS, MERS, and the endemic coronaviruses including HKU1, OC43, NL63, and 229E, and comparing these responses to those from natural infection as a function of disease severity, will be critical for understanding the immune response and ultimately delivering a safe and effective vaccine.

For effective application in vaccine clinical trials, a highly specific and highly quantitative assay is required to enable accurate quantitative assessment of antibody responses. Given the rapid timelines for vaccine development already underway, a multiplexed assay that can measure vaccine-induced antibody response to a variety of related antigens simultaneously is highly desirable for both time and cost savings. The VaxArray platform (InDevR, Inc., Boulder, CO) is a microscale, multiplexed, microarray-based immunoassay platform that has been well-validated for use in influenza vaccine antigen characterization,^21,22^ and has been adapted for serological analysis of coronaviruses with the recent availability of the Coronavirus (CoV) SeroAssay. Specifically, nine unique coronavirus spike protein antigens are printed in replicate in a microarray format, providing the ability to perform simultaneous analysis of antibody responses to all 9 antigens in a single, 2-hour assay. The 9 proteins represented on the microarray are full-length spike, receptor binding domain (RBD), and the S2 extracellular domain of SARS-CoV-2, and the spike proteins from SARS, MERS, HKU1, OC43, NL63, and 229E. In addition, the platform is antigen-sparing, requiring ~200x less antigen to manufacture than a traditional plate-based ELISA, which is particularly important during this time of strained supply chains.

This study reports on the VaxArray CoV SeroAssay linear dynamic range, limit of detection, specificity, reproducibility, accuracy, and investigates assay performance on a retrospective set of 263 blinded, de-identified human serum and plasma specimens to demonstrate positive and negative percent agreement to a mixed reference method of RT-PCR on a patient-matched specimen and collection date prior to the COVID-19 outbreak. An easy-to-use, high information content assay with the capability to evaluate antibody response to a variety of coronavirus spike proteins will aid in monitoring the immune response during COVID-19 candidate vaccine clinical trials and ultimately facilitate the delivery of a safe and effective vaccine.

## 2. MATERIALS AND METHODS

### 2.1 VaxArray Coronavirus SeroAssay Standard Procedure

The VaxArray Coronavirus SeroAssay Kit (#VXCV-5100, InDevR, Inc.) contains four microarray slides, printed with 16 replicate arrays per slide, an optimized Protein Blocking Buffer (VX-6305), Wash Buffer 1 concentrate (VX-6303), and Wash Buffer 2 concentrate (VX-6304). Prior to use, microarray slides were equilibrated to room temperature for 30 min in the provided foil pouch. Prepared standards and specimens were diluted at least 1:100 in Protein Blocking Buffer and applied to the microarray and allowed to incubate in a humidity chamber (VX-6200) on an orbital shaker at 80 rpm for 60 minutes. After incubation, samples were removed using an 8-channel pipette, and the microarray was subsequently washed by applying 50 μL of prepared Wash Buffer 1. Slides were washed for 5 minutes on an orbital shaker at 80 RPM after which the wash solution was removed via 8-channel pipette. During sample incubation, Anti-human IgG Label (VXCV-7623) and/or anti-mouse IgG Label (VXCV-7620) were prepared by first diluting the label 1:10 in PBB, and aliquoting into 8-tube PCR strips after which 50 μL of label mixture was added to each array using an 8-channel pipette. Detection label was incubated on the slides in the humidity chamber for 30 minutes before subsequent, sequential washing in Wash Buffer 1, Wash Buffer 2, 70% Ethanol, and finally ultrapure water. Slides were dried using a compressed air pump system and imaged using the VaxArray Imaging System (VX-6000).

### 2.2 Linear Dynamic Range and Lower Limit of Quantification Analysis

A study to determine the lower limit of quantification and linear dynamic range of the different capture antigens represented was executed using monoclonal antibodies that target the spike proteins of SARS-CoV-1 (MRO-1214LC, CR3022, Creative Biolabs), SARS-CoV-2 (GTX632604, Genetex), MERS (40069-MM23, Sino Biological), and HKU1 (40021-MM07, Sino Biological). The CR3022 antibody targeting SARS-CoV-1 is known to bind the the nCoV(ii) RBD antigen, the SARS antigen (and the nCoV(i) full-length spike antigen to a much weaker extent), and the SARS-CoV-2 Genetex antibody is known to bind the nCoV(i) full-length spike antigen (and the nCoV(iii) S2 antigen to a much weaker extent). The four antibodies were mixed, and a 13-point serial dilution in Protein Blocking Buffer and three blank wells containing Protein Blocking Buffer without antibody were prepared, with each sample subsequently analyzed on the VaxArray CoV SeroAssay according to the operation manual with one exception: because the anti-SARS-CoV1 antibodies are human antibodies and the other three antibodies are mouse antibodies, antibodies were detected with a mixture of anti-mouse and anti-human IgG secondary antibody labels (VXCV-7620 and VXCV-7326, InDevR, Inc., respectively). After analysis, the median signals extracted from the VaxArray Imaging System software for each relevant capture antigen for each dilution as well as for the blanks were analyzed, with the serial dilutions plotted as a function of the known concentration of the antibody and a series of moving 4-point linear fits applied to the data. The upper limits of quantification (ULOQ) were calculated for each of the relevant capture agents as the back-calculated concentration at the highest RFU signal that was within the highest 4-point fit with an R^2^ value exceeding 0.95. The lower limits of quantification (LLOQ) for each of the relevant capture antigens was calculated for each blank sample as the back-calculated concentration at the background-subtracted median signal of the blank plus 5 standard deviations of the blank. This value was then averaged over the 3 blanks. In addition, the linear dynamic range (LDR) was calculated as ULOQ/LLOQ for each relevant capture antigen.

Because monoclonal antibodies binding to the OC43, NL63, or 229E capture antigens were not available at the time of testing, linearity and limit of detection for these 3 capture antigens was explored using a limiting endpoint dilution series of a pooled human serum sample known to be positive for antibodies to all four human CoVs: OC43, NL63, HKU1, and 229E. The pooled human serum sample was used to create a 13-point serial dilution in Protein Blocking Buffer with all samples subsequently analyzed in duplicate on the VaxArray CoV SeroAssay according to the operation manual. Data was extracted in the same manner as for the mixed monoclonal antibody analysis to determine the signals at the ULOQ and LLOQ (without back-calculating to concentration based on the linear fits, as the concentration in each of the dilutions is unknown). The LDR was expressed as the signal at the ULOQ divided by the signal at the LLOQ. The limiting endpoint dilution titer was also presented as the highest dilution factor at which the signal exceeded the signal at the LLOQ.

### 2.3 Specificity

Specificity of the capture antigens was investigated using the same 4 monoclonal antibodies described for use in the LLOQ and LDR analysis. No monoclonal antibodies were available at the time of testing that target OC43, NL63, or 229E, and so specificity for these capture antigens was not assessed. However, we did assess specimens positive for all 4 endemic coronaviruses that were collected prior to the SARS-CoV-2 outbreak to examine potential cross-reactivity with any of the 3 SARS-CoV-2 antigens. A total of 132 serum samples known to be negative for the presence of antibodies to SARS-CoV-2 based on date of collection prior to December 2019, including 33 specimens from pediatric donors age 2-16, were analyzed via the standard VaxArray CoV SeroAssay procedure at a 1:100 dilution in PBB.

### 2.4 Reproducibility and Accuracy

To assess reproducibility and accuracy, a pooled human serum sample known to be positive for antibodies to SARS-CoV-2 (and known to bind all 3 SARS-CoV-2 antigens on the microarray) and all 4 of the endemic coronaviruses (HKU1, OC43, NL63, and 229E) was prepared in adequate volume to run a large number of replicates. This sample did not contain any antibody reactive to the MERS capture antigen, and therefore, reproducibility and accuracy of the assay’s ability to detect antibodies that bind to the MERS spike protein was not assessed. This study examined a single operator over three days of testing, as previous studies (data not shown) indicated little user-to-user or instrument-to-instrument variability. On days 1 and 2 of testing, a single slide containing an 8-point calibration curve (7 standards and a blank, analyzed at arbitrary relative concentrations of 1.0, 0.8, 0.6, 0.4, 0.2, 0.1, 0.05, and 0) was run alongside 8 replicates of an intermediate dilution of the same sample (expected to be at 0.4). On the 3^rd^ testing day, three additional slides of 16 replicates each were analyzed alongside the first slide, for a total of 72 replicate microarrays (the standard curve on slide 1 was used to analyze data on all 4 slides in the experiment). The above testing was performed on 3 unique slide manufacturing lots for a total of 216 replicate analyses over the 3 days and 3 lots. The quantitative mode of the VaxArray Imaging System software was used to generate back-calculated concentrations for each of the replicates analyzed for data analysis, using the relevant standard curve for each 1-slide or 4-slide analysis on each day. Averages over each set of 8 replicates (each slide) as well as over all 216 replicates were calculated and are presented as % coefficient of variation (%CV). To assess accuracy or % recovery (% of expected value), the same data were analyzed as the calculated concentration divided by the expected concentration of 0.4, expressed as a percentage. The % recovery over all 216 replicates analyzed are presented. Precision and accuracy data can be found in Table 3.

### 2.5 Clinical Sensitivity and Specificity/Positive and Negative Percent Agreement

To determine the positive (PPA) and negative (NPA) percent agreement of the VaxArray CoV SeroAssay with a known result, 263 retrospective, deidentified human specimens (260 serum, 3 plasma) were obtained from the authors’ institutions, collaborators, and commercial sources. Commercial specimen sources included Boca Biolistics (Pompano Beach, FL), US Biolab (Rockville, MD), and Lee BioSolutions (Maryland Heights, MO). Specimens from Colorado Children’s Hospital (Denver, CO) were collected under a Colorado Multiple IRB (COMIRB) approved protocol. Specimens obtained from Lankenau Institute for Medical Research (LIMR, Wynnewood, PA) were collected under a Main Line Hospitals IRB approved protocol. Deidentified specimens from Veritas, PA (Belton, TX) were obtained under an AspireIRB-approved protocol. Deidentified specimens from Mount Sinai Icanh School of Medicine (New York, NY) were obtained under a license agreement which indicates appropriate IRB approval and informed consent for the specimens provided. Additional details regarding clinical specimens included can be obtained from the authors upon request.

The reference method used for specimens positive for SARS-CoV-2 antibodies was an RT-PCR result for a donor-matched specimen. The reference method used for specimens negative for antibodies to SARS-CoV-2 was either a negative RT-PCR result for a donor-matched specimen or a known collection date prior to the COVID-19 outbreak in late 2019. All specimens obtained from Colorado Children’s Hospital were also analyzed independently by ELISA in a CLIA-certified laboratory using both an NP-based ELISA (Epitope Diagnostics, San Diego, CA) and by an S1-based ELISA (Euroimmun Ag, Germany). These ELISA results were used as orthogonal information to further investigate discrepant results. Testing personnel were blinded to these orthogonal results prior to completing VaxArray CoV SeroAssay analysis.

The sample set included 132 negatives and 131 positives, with all positives collected at least 14 days after onset of symptoms. Ninety-two (92) of the specimens were received with a known donor age. Of note, 33 of the negative specimens were from pediatric donors between the ages of 2 and 16. All specimens were internally blinded by someone not involved with the testing prior to analysis. Specimens were diluted 1:100 in PBB and carried through the VaxArray CoV SeroAssay procedure as described in the Methods section. After imaging was complete, median signal data from the VaxArray Imaging System for the sample set were exported into MS Excel and analyzed. From the median signal data, the signal to background value (S/B) was calculated for each of the 9 antigens on the chip by taking the median of 9 replicate spots for each antigen and dividing by the background of the microarray.

The diagnostic algorithm used a multi-antigen approach. Specimens were considered positive for SARS-CoV-2 antibodies by the VaxArray CoV Assay if both of the following two conditions were met: 1) the S/B on the nCoV(i) antigen exceeded the average S/B of the bank of negative specimens + 3 standard deviations of the average, AND 2) the sum of the S/B values for all 3 nCoV antigens exceeded the sum of the average S/B of the bank of negative specimens + 6 standard deviations. Once VaxArray CoV SeroAssy results were determined, PPA and NPA were calculated compared to the reference method and reported with associated 95% confidence intervals. Confidence intervals reported are two-sided Wilson Scores with no continuity correction.

## 3. RESULTS

### 3.1 Assay Principles

The VaxArray CoV SeroAssay is a multiplexed immunoassay that consists of 9 coronavirus spike proteins printed in a microarray format as schematically illustrated in Figure 1. Each of 9 antigens are printed in 9 replicate spots in a single microarray, with 16 identical microarrays printed on each slide. Details regarding the capture antigens are found in Table 1. For quantitative analysis, serum samples diluted in a blocking buffer are analyzed alongside a serial dilution of an appropriate standard material, which is utilized to quantify the antibody binding to each of the 9 antigens on the microarray. Antibodies in serum are captured by the printed antigens and are subsequently labeled with a fluorescent species-specific IgG label for detection. Spatial separation of the 9 antigens enables multiplexed analysis of antibody binding to a variety of coronavirus antigens. For qualitative analysis, serum specimens are diluted in a blocking buffer and analyzed at a single dilution factor and compared to an established cutoff value based on responses from a bank of known negative samples.

**Figure 1.**
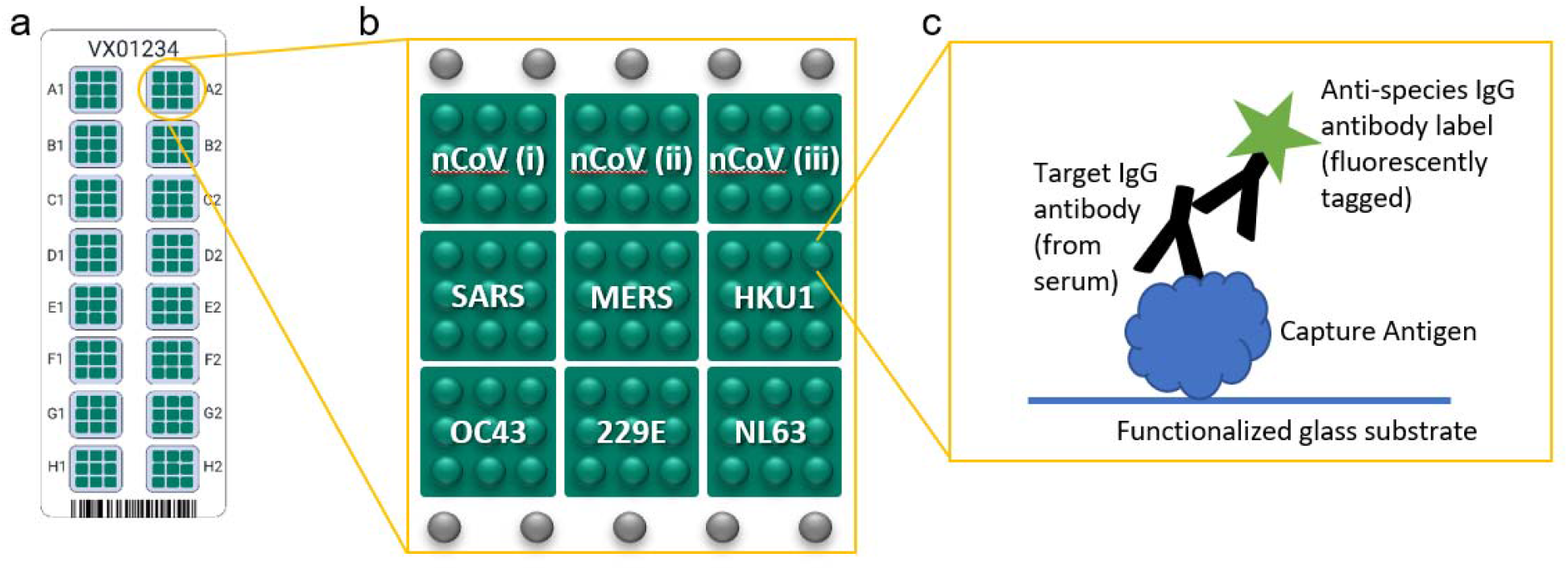
**(a)** Schematic illustration of VaxArray CoV SeroAssay slide with 16 identical microarrays labeled A1 through H2, **(b)** Layout of an individual microarray including 9 unique capture antigens labeled nCoV(i) through NL63, each printed as 9 replicate spots. Fiducial markers are shown as grey spots in rows above and below capture antigens. **(c)** Schematic of the immunoassay principle in which capture antigen binds target antibodies from serum, and target antibodies are labeled using a species-specific IgG secondary antibody label that contains a fluorescent tag.

**Table 1:** Capture Antigens.

| Name on chip | Expression System | Antigen |
| --- | --- | --- |
| nCoV(i) | Mammalian | Full length spike (S1 + S2) |
| nCoV(ii) | Mammalian | S1 Receptor Binding Domain (RBD) |
| nCoV(iii) | Insect | S2 extracellular domain |
| SARS(i) | Mammalian | S1 |
| MERS | Mammalian | S1 |
| HKU1(i) | Mammalian | S1 |
| OC43(i) | Mammalian | S1 |
| 229E(i) | Insect | Full length spike (S1 + S2) |
| NL63(i) | Mammalian | S1 |

### 3.2 VaxArray CoV SeroAssay is Quantitative with Low ng/mL Sensitivity

To demonstrate the quantitative ability of the assay, a study was executed using a mixture of mouse and human monoclonal antibodies (mAbs) that target SARS-CoV-1, SARS-CoV-2, MERS, and HKU1 (see the Methods section for details). The 4 antibodies were mixed, and a 13-point dilution was analyzed using a mixture of anti-mouse and anti-human IgG labels. Figure 2 shows representative 13-point dilutions demonstrating linearity with dilution for the nCoV(i), (ii), and (iii) antigens, respectively. Lower limits of quantification (LLOQ) for the 6 targeted antigens ranged from 0.32 ng/mL to 1.99 ng/mL, with associated linear dynamic ranges (LDR) from 76 – 911x. Because there were no available monoclonal antibodies for OC43, NL63, or 229E at the time of testing, the linearity with dilution of the assay for antibody binding to these antigens was investigated by determining a limiting endpoint dilution titer using a pooled human serum sample known to be positive for all 4 endemic human coronaviruses. Table 2 shows the calculated LLOQ, upper limit of quantification (ULOQ), and LDR for the captures for which mAbs were available, as well as the limiting endpoint dilution titers (which range from 4000 to 16000) and LDRs for OC43, NL63, and 229E (which range from 8x to 32x).

**Figure 2.**
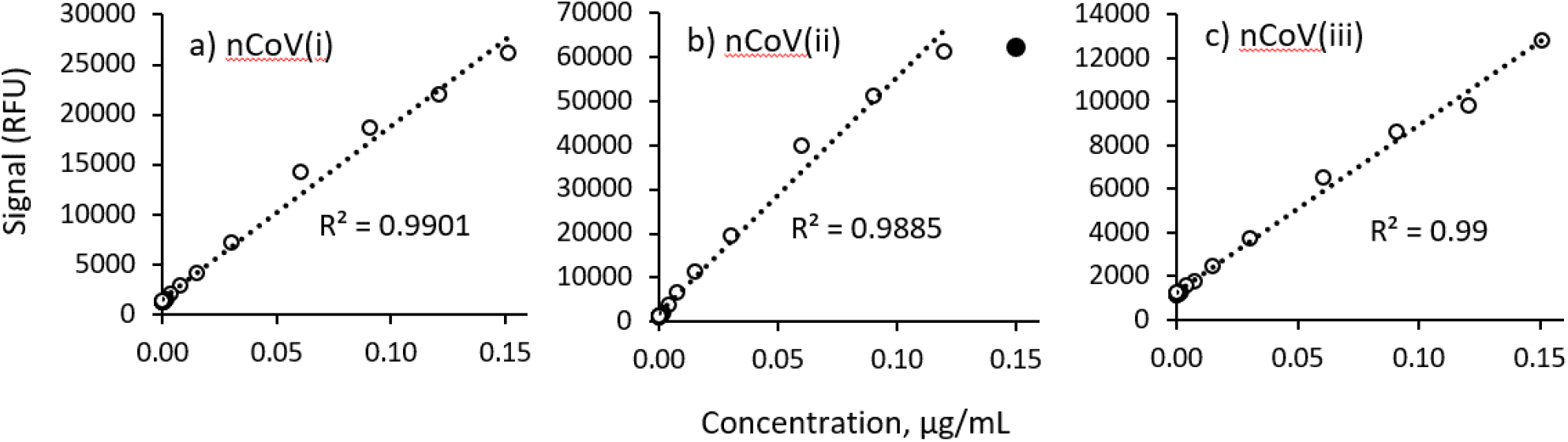
13-point dilution series of monoclonal antibodies reactive to the SARS-CoV-2 spike protein capture antigens along with associated R^2^ values for the linear regressions (n=1 for each datapoint): a) nCoV(i), b) nCoV(ii), and c) nCoV(iii). In panel b), the filled circle at the highest concentration was not included in the linear fit since it is outside the linear range.

**Table 2:** Sensitivity, Linear Dynamic Range of all 9 CoV SeroAssay Capture Antigens

| *Metrics determined with monoclonal antibodies* |  |  |  |
| --- | --- | --- | --- |
| Capture Antigen | LLOQ (ng/mL) | ULOQ (ng/mL) | LDR (no units) |
| nCoV(i) | 1.40 | 150 | 107 |
| nCoV(ii) | 0.47 | 120 | 255 |
| nCoV(iii) | 1.99 | 151 | 76 |
| SARS | 0.55 | 150 | 273 |
| MERS | 1.00 | 911 | 911 |
| HKU1 | 0.32 | 59 | 184 |
| *Metrics determined with pooled human serum* |  |  |  |
| Capture Antigen | Limiting Endpoint Dilution Titer | LDR (no units) |  |
| OC43 | 8000 | 32 |  |
| 229E | 16000 | 32 |  |
| NL63 | 4000 | 16 |  |

### 3.3 VaxArray CoV SeroAssay Demonstrates Reasonable Specificity between SARS-CoV-2 and Endemic Coronaviruses

Monoclonal antibodies reactive to SARS-CoV-2, SARS-CoV-1, MERS, and HKU1 were analyzed to investigate specificity, with representative images shown in Figure 3. No monoclonal antibodies were available at the time of testing that specifically target OC43, NL63, or 229E. One hundred thirty-two (132) human serum specimens negative for antibodies to SARS-CoV-2 were analyzed, and none showed reactivity to the nCoV(i) full-length spike or nCoV(ii) RBD antigens. Thirteen (13) specimens did show some reactivity to the nCoV(iii) (S2) antigen, indicating some degree of non-specificity. Thirty-three (33) of the 132 negative specimens were pediatric samples from donors aged 2-16, one of which was cross-reactive on the nCoV(iii) S2 antigen. Representative microarray images of human serum samples negative for SARS-CoV-2 with different reactivities to the endemic CoV capture antigens on the microarray are shown in Figure 4 (panels b through h) along with the associated donor ages; also in Figure 4 (panels i through l) are representative images of human serum samples from donors of unknown age known to be positive for SARS-CoV-2 antibodies for comparison. A reminder of the microarray layout is included as Figure 4 panel a for clarity. In addition, for donor-matched serum from SARS-CoV-2 RT-PCR positive patients, we do observe some cross-reactivity of SARS-CoV-2 antibodies with the SARS-CoV-1 antigen.

**Figure 3.**
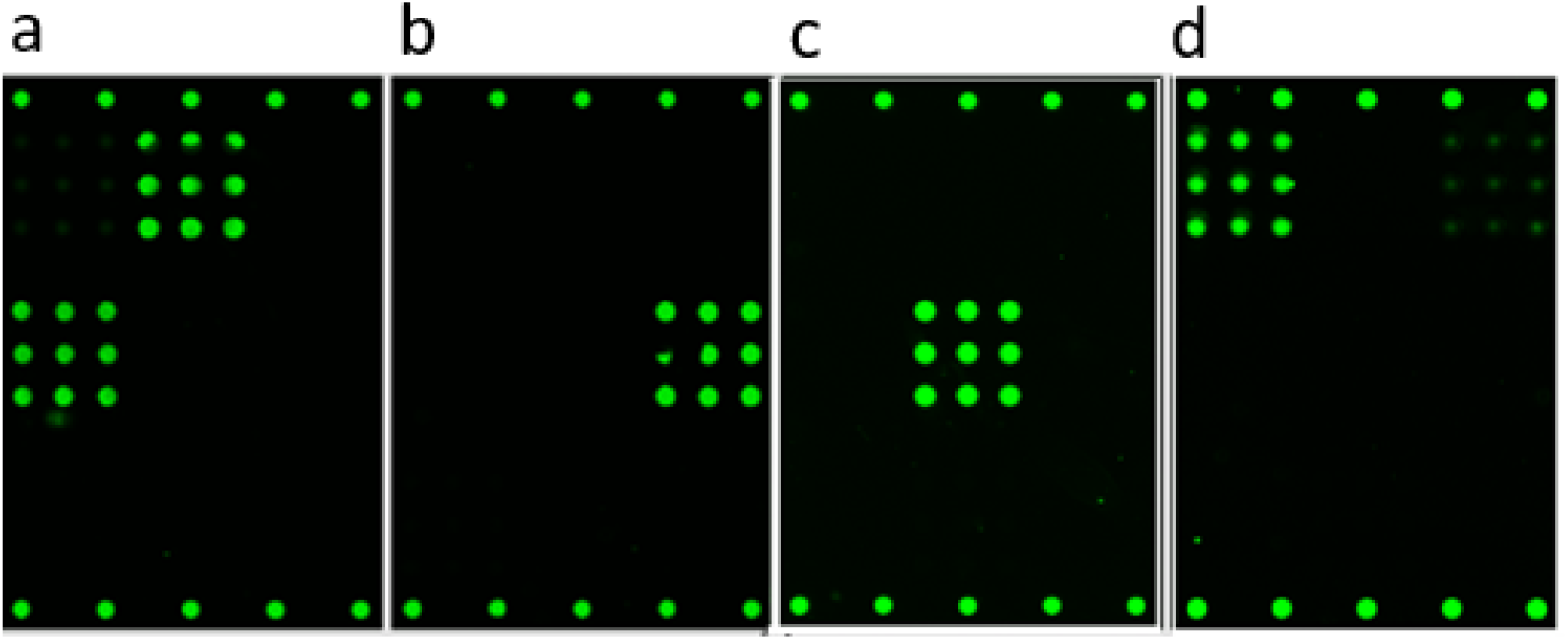
Fluorescence microarray images illustrating binding of monoclonal antibodies to the CoV SeroAssay. (a) CR3022 SARS-CoV-1 antibody from Creative Biolabs binding to the nCoV(ii) and SARS antigens, (b) 40021-MM07 HKU1 antibody from Sino Biological binding to HKU1 antigen, (c) 40069-MM23 MERS antibody from Sino Biological binding to the MERS antigen, and (d) GTX632604 SARS-CoV-2 antibody from Genetex binding to the nCoV(i) and nCoV(iii) antigens.

**Figure 4.**
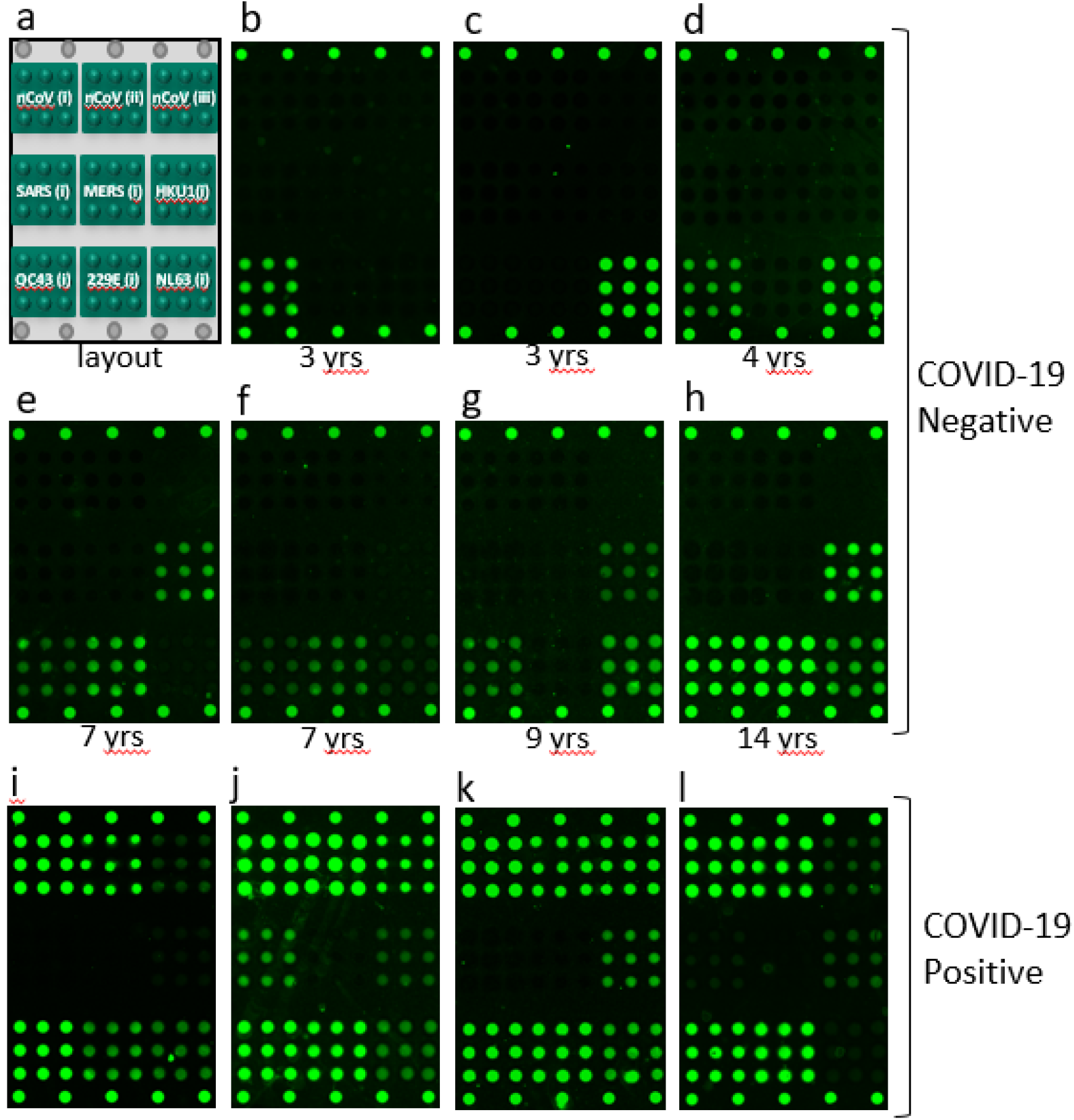
(a) Schematic illustration of microarray layout, and representative fluorescence images of the VaxArray CoV SeroAssay microarray in (b) through (l). Images (b) through (h) show the variety of responses to the endemic human CoV capture antigens for specimens from pediatric donors (ages noted for each image), and images (i) through (l) show responses from 4 unique COVID-19 positive donors of unknown age.

### 3.4 VaxArray CoV SeroAssay shows 11% Overall %CV for Replicate Measurements

To assess reproducibility and accuracy, a pooled human serum sample positive for antibodies to SARS-CoV-2, and also known to be reactive to the SARS antigen and the 4 endemic coronaviruses was prepared (no reactivity to MERS antigen), and used to create a serial dilution to be used as a standard curve as well as replicate aliquots of an intermediate dilution to be analyzed in replicate. Seventy-two (72) replicates of the intermediate dilution were tested by a single operator over 3 days on each of 3 manufacturing lots of microarray slides, for a total of 216 replicate analyses. Previous studies indicated little user-to-user or instrument-to-instrument variability (data not shown). The replicates analyzed on each day were quantified in arbitrary concentration units using the day- and lot-specific calibration curve, with the highest concentration of the serum sample arbitrarily assigned a concentration of 1. Replicates were run at an expected concentration of 0.4.

Table 3 shows the % CV in the back-calculated concentration value obtained on each relevant capture antigen for all 216 replicate measurements over all 3 days and all three lots of slides, with values ranging from 7 to 19 %CV for the 9 antigens. In addition, the %CV of the 8 replicates run on each slide in the study was analyzed to assess the intra-slide precision, resulting in an average intra-slide CV that ranged from 5% - 8% for each of the 9 antigens, for an overall intra-slide CV of 6%.

**Table 3:** Precision, Accuracy of all 9 CoV SeroAssay Capture Antigens*

| Capture Antigen | Average Accuracy, with associated % CV | Average Precision, expressed as % CV |
| --- | --- | --- |
| nCoV(i) | 94% (9%) | 10% |
| nCoV(ii) | 97% (10%) | 11% |
| nCoV(iii) | 88% (17%) | 19% |
| SARS(i) | 92% (10%) | 11% |
| HKU1(i) | 90% (9%) | 10% |
| OC43(i) | 96% (9%) | 10% |
| 229E(i) | 95% (7%) | 7% |
| NL63(i) | 91% (10%) | 11% |

\*Data represents n=216 pooled serum samples analyzed (single user, 3 slide lots over 3 days). MERS capture antigen was not evaluated in this study.

### 3.5 VaxArray CoV SeroAssay Accuracy (% Recovery) Ranges from 88% to 97%

The data generated for the reproducibility study were also used to assess accuracy expressed as % of expected result (% recovery). The calibration curves generated using the quantitative mode of the VaxArray Imaging System software were utilized to determine the relative concentration present for all the replicates for each of the relevant antigens. The concentrations determined were then averaged for all 216 replicates and compared to the expected concentration. These accuracy data are presented in Table 3 and range from 88 to 97% for the various capture antigens for an overall average accuracy of 92.5%.

### 3.6 VaxArray CoV SeroAssay demonstrated PPA (Sensitivity) of 98.5% and NPA (Specificity) of 100%

Two hundred sixty-three (263) deidentified specimens (260 serum, 3 plasma) were received for testing. The sample set included 132 specimens known to be negative for COVID-19 by RT-PCR of a donor-matched specimen or were collected prior to the COVID-19 outbreak in late 2019, and 131 specimens known to be positive for COVID-19 by RT-PCR of a donor-matched specimen. All serum specimens from COVID-19 positive donors were collected at least 14 days after the initial onset of symptoms, and representative fluorescence microarray images from positive specimens are shown in Figure 4. This study resulted in positive percent agreement (PPA) of 98.5% (95% CI of 94.6 – 99.6 95%, Wilson Score, two-sided with no continuity correction) and negative percent agreement (NPA) of 100% (95% CI of 97.2 – 100%, Wilson Score, two-sided with no continuity correction) with the mixed reference method described. This dataset resulted in zero false positives and 2 false negatives by the VaxArray CoV SeroAssay.

To demonstrate the range of underlying quantitative responses obtained from these clinical specimens, Figure 5 shows a scatter plot of signal to background ratios obtained for the nCoV(i) capture antigen for the 131 specimens from donors expected to be positive by RT-PCR, with signal to background ratios sorted from low to high. The data show that the antibody responses of these specimens from COVID-19 positive donors are highly variable.

**Figure 5.**
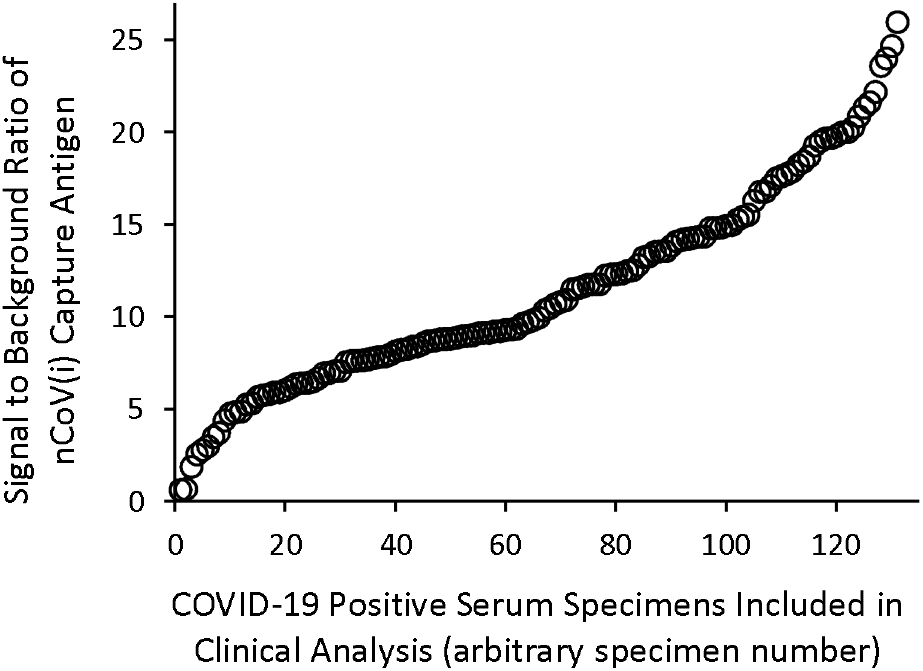
Signal to background ratio of the nCoV(i) antigen for the 131 human serum specimens included in the clinical analysis that were from donors positive for SARS-CoV-2 by RT-PCR, sorted from lowest to highest signal to background ratio.

## 4. DISCUSSION

The availability of an accurate, precise, sensitive and specific multiplexed antibody quantification assay for COVID-19 vaccine development is an important tool for vaccine manufacturers to enhance and speed up pre-clinical and clinical trials and to ultimately aid in the delivery of a safe and effective vaccine. Much of the development of antibody-based tests for COVID-19 has focused on the development of qualitative ELISA and lateral flow-type tests for applications in diagnostics and seroprevalence to assess an individual’s antibody response post-infection. In contrast, this assay was specifically developed to provide information regarding antibody response for the wide variety of almost 200 COVID-19 vaccines currently under development.^13,14^ The VaxArray CoV SeroAssay capture antigens represent not only different forms of the spike protein from SARS-CoV-2, including full-length spike, receptor binding domain (RBD), and the S2 extracellular domain, but also includes spike proteins from a variety of other endemic and potentially pandemic coronaviruses as outlined in Table 1 to enable comprehensive analysis of the polyclonal antibody responses produced post-vaccination or post-infection.

The VaxArray CoV SeroAssay is quantitative with a linear response over a wide range of concentration. In addition, further dilution of the starting specimen can easily enable samples above the linear dynamic range to be effectively quantified. It is difficult to compare analytical sensitivity of the VaxArray CoV SeroAssay to the wide variety of available IgG ELISA assays for measuring antibodies to SARS-CoV-2 based on published data, as most of these assays are qualitative and are not intended for quantitative analysis. As a result, performance metrics for other IgG immunoassays focus on clinical sensitivity and specificity and typically do not report analytical sensitivity. Limits of quantification of the VaxArray CoV SeroAssay are comparable to other fluorescence- and chemiluminescence-based immunoassays that are typically in the sub- to low-ng/mL range.^23^ This sensitivity is clearly adequate for measuring antibody responses in human serum specimens after natural infection, as evidenced by the data summarized in Table 4, in which all 263 serum samples were diluted 100-fold in Protein Blocking Buffer prior to analysis. For the 131 known positive specimens in this dataset, 80% of the specimens had a signal close to fluorescence saturation on the nCoV(i) capture sequence (a signal exceeding 60000 RFU with fluorescence saturation occurring at 65535 RFU).

**Table 4:**
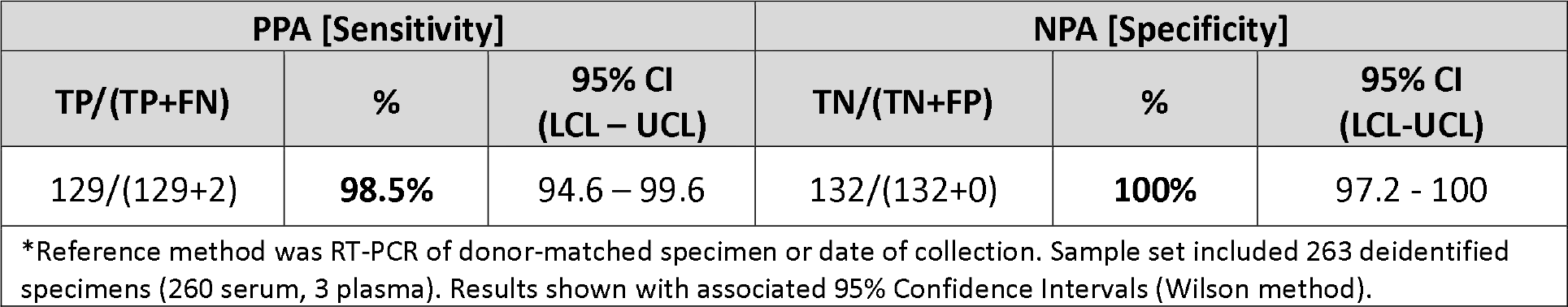
Positive and Negative Percent Agreement between VaxArray CoV SeroAssay for the detection of SARS-CoV-2 compared to a Mixed Reference Method*

| PPA [Sensitivity] |  |  | NPA [Specificity] |  |  |
| --- | --- | --- | --- | --- | --- |
| TP/(TP+FN) | % | 95% CI<br>(LCL – UCL) | TN/(TN+FP) | % | 95% CI<br>(LCL-UCL) |
| 129/(129+2) | **98.5%** | 94.6 – 99.6 | 132/(132+0) | **100%** | 97.2 - 100 |
| *Reference method was RT-PCR of donor-matched specimen or date of collection. Sample set included 263 deidentified specimens (260 serum, 3 plasma). Results shown with associated 95% Confidence Intervals (Wilson method). |  |  |  |  |  |

Using monoclonal antibodies, we demonstrated the VaxArray CoV SeroAssay shows the expected specific response for 6 of the 9 antigens, with the monoclonal antibody to SARS-CoV-1 (CR3022) producing signal on both the nCoV(ii) RBD antigen and the SARS antigen (SARS-CoV-1 spike protein). This is not surprising given that SARS-CoV-1 and SARS-CoV-2 share a cellular receptor and have significant sequence homology [Ye]. The SARS-CoV-2 monoclonal antibody (Genetex) bound to both the nCoV(i) full-length spike protein and nCoV(iii) S2 extracellular domain. Importantly, though, neither of these antibodies bound to any of the human endemic coronavirus antigens on the array. As there were no specimens available from donors previously infected with COVID-19 but known to be negative for previous infections with all 4 endemic coronaviruses (likely due to the very high proportion of seropositivity to some or all of the endemic CoVs in the human population),^24^ examining specificity of the polyclonal antibody response in humans in this manner is difficult. However, in our analysis of 132 human serum specimens from COVID-19 negative donors, no samples produced a signal to background value exceeding the threshold for the nCoV(i) or nCoV(ii) antigens. Thirteen (13) samples produced a signal to background value exceeding the threshold for nCoV(iii), indicating some cross-reactivity of COVID-19 negative human serum on this antigen. Only one of the 13 specimens from COVID-19 negative donors that produced signal on nCoV(iii) was from a pediatric patient. In further analyzing pediatric specimens from COVID-19 negative donors, we note that a variety of responses to the endemic CoVs are present as shown in Figure 4. As expected, younger children in general tend to have antibodies that bind to some of the 4 endemic coronaviruses, whereas older children and adolescents often show antibodies to all 4 in most cases. This is consistent with reports that seroconversion increases with age, with most adults showing seroconversion to all 4 human CoVs.^24^

Importantly, in a pre-clinical or clinical trial for a candidate COVID-19 vaccine, specimens can be analyzed prior to vaccine administration to determine a baseline response to each of the antigens on the array. Therefore, serum reactivity to the nCoV(iii) antigen pre-vaccination can be accounted for by this baseline response. This response can then be compared to the response as a function of time post-vaccination, and any changes in reactivity to all 9 antigens can be quantitatively assessed to provide a broader profile of the antibodies produced after vaccination. That this can be accomplished in a single test with a 2-hour turnaround time means that serology analysis during a clinical trial can be completed in less time and for less cost.

In a set of experiments involving 216 replicate analyses, the VaxArray CoV SeroAssay demonstrated good precision and accuracy, as shown in Table 3. The data represented a single user testing over 3 days and on 3 unique lots of microarray slides (a total of 6 unique slides per lot). Within a single slide of 8 replicates, the average % CV of the measurements was 6%, indicating excellent precision on a single slide. This %CV ranged from 7% to 19% for the 9 antigens, averaging over all 3 days and all 3 lots. A higher %CV representing day-to-day and lot-to-lot variation is reasonable given the variables represented. As shown in Table 3, nCoV(iii) demonstrated the highest variation. This was due to a single lot of microarray slides (lot 1) that produced a higher than expected 25% CV variation for this antigen, whereas lots 2 and 3 produced only 10% and 13% CV, respectively. Interestingly, however, the other 7 capture antigens investigated in this study did not experience a higher %CV for lot 1. This single lot may have experienced a printing artifact for this antigen, and the absolute signals generated on this nCoV(iii) antigen for this study were only ~2x LLOQ. Regardless of the root cause, printing optimization efforts are underway to improve the lot-to-lot consistency in performance of this antigen. Accuracy expressed as % recovery (% of expected result) was also quite good, ranging from 88% to 97% over the entire dataset of n=216 replicates. Unsurprisingly, nCoV(iii) also suffered from the lowest accuracy on the same lot of slides showing lower than expected precision. The average accuracy of the nCoV(iii) result was 80%, 85%, and 100% for lots 1, 2, and 3, respectively.

The clinical specimen analysis summarized in Table 4 indicates the VaxArray CoV SeroAssay has excellent positive and negative percent agreement compared to a mixed reference method of RT-PCR status for a matched donor specimen or collection date prior to the COVID-19 outbreak in late 2019. The cutoff established for the VaxArray CoV SeroAssay takes advantage of the unique multiplexed capability of the assay by using a multi-antigen approach to thresholding to increase the confidence in a positive or negative call. Specifically, the signal on the nCoV(i) antigen was used as a first ‘gate’ to a positive call, and the sum of the three nCoV antigens was used as a secondary ‘gate’ to a positive call. Both cutoffs had to be exceeded in order to make a positive call, providing additional confidence against false positive results. This thresholding methodology resulted in 98.5% positive agreement (129/131) and 100% negative agreement (132/132) with the mixed reference method. The two specimens that produced false negative results by the VaxArray CoV SeroAssay were obtained from Children’s Hospital of Colorado, and both were independently found to be negative by two alternative IgG-based ELISA assays, and both had very low titers by a virus neutralization assay (data not shown). All orthogonal analyses were conducted by Children’s Hospital of Colorado, and InDevR staff analyzing the clinical specimens by the VaxArray CoV SeroAssay were blinded to these results. These additional serological analyses indicate concordance with the VaxArray CoV SeroAssay results, likely indicating that either the donors produced little to no antibody response after infection, or that the associated RT-PCR results were false positives.

To highlight the underlying quantitative response generated for these clinical serum specimens, Figure 5 shows the signal to background ratio produced on the nCoV(i) capture antigen for all 131 serum specimens from patients known to be COVID-19 positive by RT-PCR, sorted from lowest to highest. Given that all specimens were analyzed at the same 1:100 dilution and the analytical data indicating the quantitative ability of the assay, these data are indicative of relative SARS-CoV-2 antibody concentrations. The two clinical specimens that produced false negative results described above are the first two datapoints in the lower left closest to the origin, with corresponding signal to background ratios of 0.63 and 0.68. The remainder of these data show that a wide range of antibody responses were observed in the positive specimens, highlighting the quantitative capabilities of the assay for assessing antibody response in COVID-19 vaccine pre-clinical and clinical trials. In addition, tools such as the VaxArray CoV SeroAssay could easily be used to correlate severity of disease with antibody titer produced, and for a wide variety of other SARS-CoV-2 to add to our current understanding.

## 5. CONCLUSION

Developers and manufacturers of candidate COVID-19 vaccines face the daunting challenge of bringing a safe and effective vaccine to market in record time to put a halt to the current global pandemic. As such, tools that empower developers and manufacturers to conduct vaccine clinical trials efficiently and to obtain the maximum amount of information in a rapid turnaround time are critical. The collective studies presented herein demonstrate that the VaxArray CoV SeroAssay is one of those tools, offering excellent analytical performance in terms of limits of quantification, high precision and accuracy and high clinical sensitivity and specificity. While most of the data herein demonstrated applicability to measurement of IgG antibodies in human serum, applicability to other animal models such as mouse or non-human primates is readily enabled using alternative anti-species label antibodies. We hope this tool will be utilized to maximize information content in the critical effort of delivering a safe and effective SARS-CoV-2 vaccine in record time.

## Data Availability

All data described in the study is available from the corresponding author.

## ACKNOWLEDGEMENTS

We acknowledge specimens received under a materials transfer agreement from Dr. Scott Dessain at Lankenau Institute for Medical Research (Wynnewood, PA), specimens received from Mount Sinai School of Medicine under a license agreement, and specimens from Dr. Aleta Bonner at Veritas, PA (Belton, TX).

## DECLARATIONS OF INTEREST

E. Dawson, K. Rowlen, and L. Kuck are stockholders of InDevR, Inc. E. Dawson, K. Rowlen, R. Blair, A. Taylor, and E. Toth are employed by InDevR Inc. L. Kuck was employed by InDevR at the time the work described was conducted. V Knight has no declarations of interest.

## AUTHOR CONTRIBUTIONS

**Erica Dawson:** project administration, supervision, methodology, resources, formal analysis, writing-original draft, visualization **Kathy Rowlen:** conceptualization, project administration, resources, writing-review and editing **Laura Kuck:** methodology, investigation, validation **Rebecca Blair:** conceptualization, methodology, resources, writing-review and editing **Amber Taylor:** supervision, methodology, formal analysis, visualization, writing-review and editing **Evan Toth:** investigation, writing-review and editing **Vijaya Knight:** resources, writing-review and editing

This research did not receive any specific grant from funding agencies in the public, commercial, or not-for-profit sectors.

## Notes

### Competing Interest Statement

E. Dawson, K. Rowlen, and L. Kuck are employed by and hold stock in InDevR, Inc. R. Blair, A. Taylor, and E. Toth are employees of InDevR.

### Funding Statement

No external funding was received for the work described herein.

### Author Declarations

All specimens were tested at InDevR, and were received completely deidentified. Specimens from Colorado Childrens Hospital (Denver, CO) were collected under a Colorado Multiple IRB (COMIRB) approved protocol. Specimens obtained from Lankenau Institute for Medical Research (LIMR, Wynnewood, PA) were collected under a Main Line Hospitals IRB approved protocol. Deidentified specimens from Veritas, PA (Belton, TX) were obtained under an AspireIRB approved protocol. Deidentified specimens from Mount Sinai Icanh School of Medicine (New York, NY) were obtained under a license agreement which indicates appropriate IRB approval and informed consent was obtained for the specimens provided.

